# WeChat, a Chinese social media, may early detect the SARS-CoV-2 outbreak in 2019

**DOI:** 10.1101/2020.02.24.20026682

**Authors:** Wenjun Wang, Yikai Wang, Xin Zhang, Yaping Li, Xiaoli Jia, Shuangsuo Dang

**Author notes:** Correspondence: Shuangsuo Dang, Department of Infectious Diseases, Second Affiliated Hospital of Xi’an Jiaotong University, 157 Xiwu Road, Xi’an, China.

## Abstract

We plotted daily data on the frequencies of keywords related to severe acute respiratory syndrome coronavirus 2 (SARS-CoV-2) from WeChat, a Chinese social media. Using “Feidian”, Chinese abbreviation for SARS, may detect the SARS-CoV-2 outbreak in 2019 two weeks earlier. WeChat offered a new approach to early detect disease outbreaks.

## Introduction

An outbreak of pneumonia of unknown cause in Wuhan, the capital of Hubei province of China, occurred in December 2019[1]. Shortly, the cause was identified as a novel coronavirus[2], which resembles severe acute respiratory syndrome coronavirus (SARS-Cov) and now is named SARS-Cov-2[3, 4]. As of February 23, 2020, 78,811 laboratory-confirmed cases and 2,462 deaths have been reported globally and the vast majority of the cases and deaths has happened in China[5]. With a series of measures such as knockdown of 13 cities in Hubei province and stringent travel restrictions throughout the nation, the disease spread is slowing down. However, China still reports quite a few new cases and new deaths every day and the epidemic may rebound as people return to workplace after extended Spring Festival holiday. Meanwhile, the number of infected patients is increasing in other countries (especially in South Korea, Japan, and Italy)[5]. Better early warning and response systems should be established to avoid such disasters.

Traditional surveillance systems, including those used by Chinese Centers for Disease Control and Prevention (CDC), typically rely on clinical, virological, and microbiological data submitted by physicians and laboratories. Due to time and resource constraints, a lack of operational knowledge of reporting systems, and regulations in these systems, substantial lags between an outbreak event and its report are common[6]. On average, the reporting delay is around two weeks for traditional surveillance systems[6, 7].

With the popularization of Internet and smartphones, an increasing number of people use social media, such as Twitter and Facebook, to share information. An event may have been posted in social media for several days or even months before its report through health institutions and official reporting structures. Internet-based search engines are an important source for health information for people from all walks of life. Analyzing data on search behaviors in turn provides a new approach for detection and monitoring of diseases and symptoms. Technologies using social media, search queries and other Internet resources offer novel and economic approaches for detecting and tracking emerging diseases and have been successfully used in the cases of SARS[8], influenza[9], dengue[10], etc. Herein, we describe the use of such an approach with WeChat, a Chinese social media, to early detect the SARS-Cov-2 outbreak in China, 2019. Internet search queries from Hubei province were also explored.

## Methods

WeChat (called Weixin in China), provided by Tencent Inc., is the largest social media in China with over 1 billion monthly active users. WeChat Index, accessed in WeChat app, is a publicly available data service that shows how frequent a specific keyword has appeared in posts, subscriptions, and search on WeChat over a period of last 90 days. Using WeChat Index, we obtained daily data from Nov 17, 2019 to Feb 14, 2020 for the keywords related to the SARS-Cov-2 disease such as “SARS”, “Feidian (Chinese abbreviation for severe acute respiratory syndrome)”, “pneumonia”, “fever”, “cough”, “shortness of breath”, “dyspnea”, “fatigue”, “stuffy nose”, “runny nose”, “diarrhea”, “coronavirus”, “novel coronavirus”, and “infection” (see the raw data in Appendix A). All the keywords except “SARS” were used in Chinese.

Baidu is the dominant Chinese Internet search engine. Baidu Index (https://index.baidu.com), provided by Baidu Inc., can show how frequent a keyword has been queried over a time period from a region. The keywords related to the SARS-Cov-2 disease as mentioned above were also explored through Baidu Index for Hubei province.

The daily data were plotted according to time for each of the keywords. As the outbreak is an isolated rather than recurrent event and the cut-off value to detect an outbreak based on social media and online search behavior is unknown, statistical analysis were not performed. The outbreak was announced by Wuhan Health Commission (WHC) on Dec 31, 2019 (D-day) and China CDC involved in investigation and response this day[11]. If WeChat Index for a keyword spiked or increased before D-day, the index for the keyword was considered as a potential candidate for the outbreak sign[12].

## Results

During the period from Nov 17, 2019 to Dec 30, 2019 (44 days), WeChat Index spiked or increased for the keywords Feidian, SARS, coronavirus, novel coronavirus, shortness of breath, dyspnea, and diarrhea (Table 1).

**Table 1:** Keywords for which WeChat Index spiked or increased during the period from Nov 17, 2019 to Dec 30, 2019.

| Keyword | A-day | The time from A-day to D-day |
| --- | --- | --- |
| Feidian | Dec 15, 2019 | 16 days |
| SARS | Dec 01, 2019 | 30 days |
| Coronavirus | Dec 30, 2019 | 1 day |
| Novel coronavirus | Dec 11, 2019 | 20 days |
| Shortness of breath | Dec 22, 2019 | 9 days |
| Dyspnea | Dec 22, 2019 | 9 days |
| Diarrhea | Dec 18, 2019 | 13 days |

A-day: the day when WeChat index for a keyword spiked or began to increase during the period from Nov 17, 2019 to Dec 30, 2019.
D-day: Dec 31, 2019, the day that the SARS-Cov-2 outbreak was announced by Wuhan Health Commission and Chinese Center for Disease Control and Prevention involved in investigation and response.

WeChat Index for Feidian stayed at a low level with small variance before Dec 15, 2019, on which it increased significantly. Then, the index persisted at relative high levels from the next day to Dec 29, 2019 and rose rapidly just on the day before D-day, up to a peak on D-day (Figure 1). The index for SARS was stable except in the first three days in December with a peak on Dec 1, 2019, on which the first known patient with laboratory-confirmed SARS-Cov-2 infection had symptom onset (Figure 1). WeChat Index for novel coronavirus had a spike on Dec 11, 2019 (Appendix B). WeChat Index for coronavirus had been roughly stable till Dec 30, 2019, when it rose rapidly (Appendix B). WeChat Index for shortness of breath as well as dyspnea began to increase from Dec 22, 2019 (Appendix B). WeChat Index for diarrhea spiked on Dec 18, 2019 (Appendix B).

**Figure 1.**
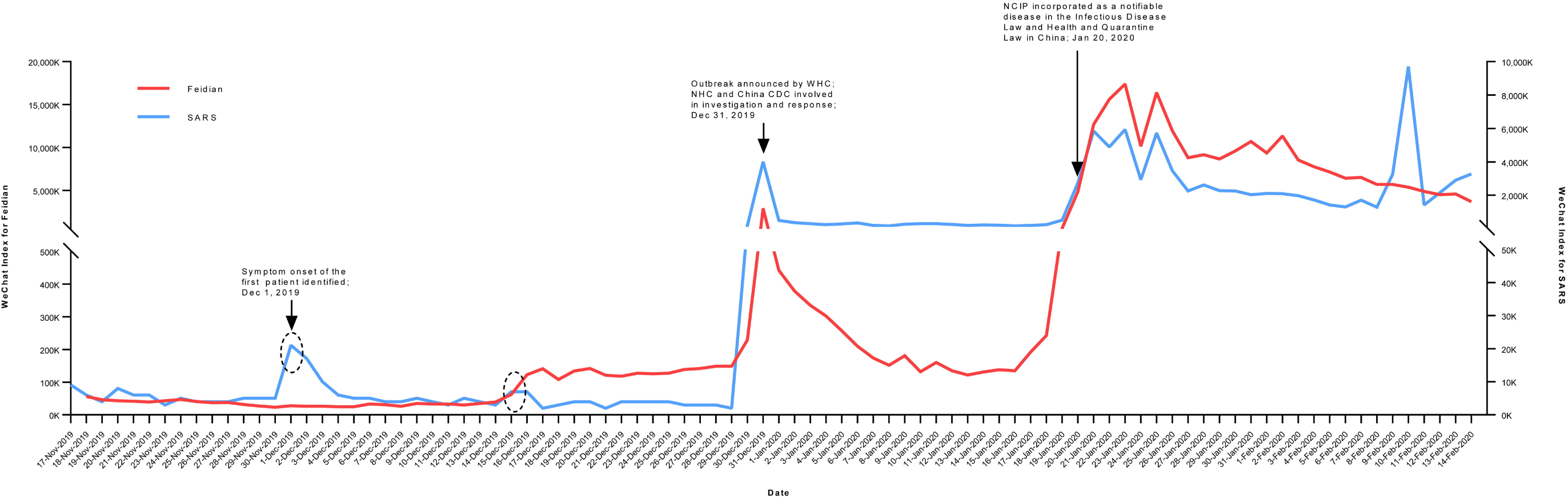
WeChat Index for Feidian and SARS (from Nov 17, 2019 to Feb 14, 2020). The index for Feidian began to rise on Dec 15, 2019 (dashed circle), persisted at relative high levels till Dec 29, 2019 and rose rapidly on Dec 29, 2019 with a peak on Dec 30, 2019. The index for SARS behaved abnormally in the first three days in December with a peak on Dec 1, 2019 (dashed circle). China CDC: Chinese Centers for Disease Control and Prevention; Feidian: Chinese abbreviation for severe acute respiratory syndrome; NCIP: novel coronavirus-infected pneumonia; NHC: National Health Commission of the People’s Republic of China; SARS: severe acute respiratory syndrome.

WeChat Index for pneumonia had spikes but showed a periodic pattern with a 7-day cycle without obvious long-term trend so that was not considered as a potential candidate for the outbreak sign (Appendix B).

Baidu Index for Feidian, SARS, pneumonia, and coronavirus rose rapidly on Dec 30, 2019, the day just before D-day. For other keywords Baidu Index showed no obvious increase during the period from Nov 17, 2019 to Dec 30, 2019 (Appendix C).

## Discussion

By exploring daily data from WeChat, a Chinese social media, we found that the frequencies of several keywords related to the SARS-Cov-2 disease behaved abnormally during a period ahead of the outbreak in China, 2019. Of these keywords, Feidian is especially worthy of attention. In 2003, the SARS outbreak caused mass panic among people in China and approximately half of the victims were health care workers[13]. Since then, Chinese physicians are alert to SARS and similar diseases[14]. If the manifestations and chest images indicate viral pneumonia and similar cases occurs in a region during a short time, they naturally think of SARS that is called Feidian in Chinese language. When suspected cases admitted to hospitals, involved physicians may mention Feidian and communicate in WeChat using this word. As showed in the current study, the activity of Feidian in WeChat began to rise on Dec 15, 2019. According to publications of early cases with laboratory-confirmed SARS-Cov-2 infection, five to eleven patients had symptom onset by this day, with the first onset on Dec 1, 2019[1, 11]. Furthermore, the activity of Feidian persisted at levels higher than those ahead of Dec 15, 2019 and it dramatically rose to a peak on the day of the outbreak announcement. Altogether, WeChat Index for Feidian is a strong warning sign for the SARS-Cov-2 outbreak. Using it may early detect the outbreak ahead of two weeks compared with the traditional surveillance systems in China.

The activity of SARS in WeChat was abnormally high in Dec 1 to 3, 2019 compared to the days ahead and after. Data by now shows that the symptom onset date of the first patient identified was Dec 1, 2019. Whether there was an inner link between the word activity in WeChat and early patients is unknown. The activity of novel coronavirus in WeChat was abnormally high on Dec 11, 2019 with an Index value of 400. However, its baseline value of WeChat Index (value equals 0 or 50) was very low compared with those of other keywords (Appendix A). It is unclear whether the abnormality was a signal or a noise. As for keywords related to symptoms such as shortness of breath, dyspnea, and diarrhea, WeChat Index increased in value ahead of 9-13 days of the outbreak announcement. These symptoms are not specific to SARS-Cov-2 infection. The increases may have some association with emergence of the disease, or just a coincidence. Though lack inner link and not performed that well in detecting the SARS-CoV-2 outbreak compared with keyword Feidian, other keywords explored in the study, including those not considered as potential candidates for the outbreak sign, and even keywords not explored in the study (such as the names of drugs treating SARS-CoV-2 infection), may still have value in future outbreak detection and monitoring. Experience from Google Flu Trends showed that a combination of potential keywords other than a single keyword is more successful[9].

Gathering and analyzing data from social media, Internet search queries, news wires and web sites represents a novel approach to early warning and detection of disease outbreaks and is a supplementary to traditional surveillance systems[6]. Such a tool, the Global Public Health Intelligence Network (GPHIN), identified the early SARS outbreak in China in 2003 more than two months earlier and first alerted the outbreak of Middle East Respiratory Syndrome Coronavirus (known as MERS-CoV) in 2012[8]. As far as we know, GPHIN and other established tools do not gather data from WeChat, the dominant Chinese social media. The current study shows that gathering and analyzing data from WeChat may be amazing to early detect disease outbreak. This may have an advantage especially for detecting outbreaks in China considering the population of WeChat users. Previous studies used search query data, including Baidu Index, to monitor the activity of emerging infectious diseases[15-17]. As far as we know, this is the first time to use WeChat data in this field. As the current study indicated, using WeChat data may perform better than using Baidu search query data in early detecting the outbreak of a new disease because people may communicate first in WeChat in the times of WeChat as a lifestyle of Chinese people[18].

The main limitation of the study lies on its retrospective nature. The outbreak is a solitary case. Using WeChat data to early detect outbreaks like this should be valued in the future. In addition, history data of WeChat Index 90 days ago is unavailable and how the index is calculated is not open.

In summary, using data of WeChat, a Chinese social media, may early detect the SARS-CoV-2 outbreak in China in 2019. If WeChat Index for the keyword Feidian were monitored, the outbreak would be detected 2 weeks earlier. Future studies can prospectively gather and analyze data from WeChat to early detect SARS-CoV-2-like outbreaks as well as outbreaks of other diseases in China. Tracing the source of keywords that behave abnormally in frequencies in WeChat, following rapid response, may become a promising approach to control a disease outbreak in its very early stage.

## Data Availability

All data are available through supplemental files.

**Appendix A. Raw data of WeChat Index for keywords related to SARS-CoV-2**.

**Appendix B. WeChat Index curves for keywords related to SARS-CoV-2 except Feidian and SARS**.

**Appendix C. Baidu Index curves for keywords related to SARS-CoV-2**.

## Notes

### Competing Interest Statement

The authors have declared no competing interest.

### Funding Statement

This work was supported by Science and Technology Research and Development Program of Shaanxi Province (Grant No. S2020-YF-YBSF-1279)

